# Predicting long-term waning of anti-spike antibody-dependent protection against COVID-19 across diverse SARS-CoV-2 immune histories

**DOI:** 10.1101/2025.11.14.25340279

**Authors:** Koki Numakura, Sho Miyamoto, Akiko Sataka, Haruko Takeyama, Tadaki Suzuki

**Author notes:** These authors contributed equally to this article (Koki Numakura, Sho Miyamoto). **Corresponding Author:** Tadaki Suzuki, Department of Infectious Disease Pathology National Institute of Infectious Diseases Japan Institute for Health Security, Toyama 1-23-1, Shinjuku, Tokyo 162-8640, Japan.

## Abstract

SARS-CoV-2 mRNA vaccines elicit robust anti-spike (anti-S) and neutralizing antibody responses; however, their kinetics beyond one year and variation across complex combination of vaccination and infection histories remain unclear. We analyzed data from 25,800 Japanese adults from four nationwide sero-epidemiological surveys (December 2021 to March 2023). Using Bayesian hierarchical models, we characterized long-term antibody trajectories stratified by vaccine doses (2–5), infection history (uninfected, pre-Omicron, Omicron), and demographics. In uninfected participants, 3–5 vaccine doses induced higher peak anti-S titers and slower waning than two doses. Notably, a history of Omicron infection, but not a history of an Omicron-adapted bivalent booster vaccination, was associated with sustained high anti-S and Omicron BA.5-neutralizing titers for over two years, irrespective of the number of vaccine doses. We then integrated these antibody-waning models with a correlates-of-protection model to predict the long-term decline in protection against symptomatic infection, stratified by specific immune history. These models provide a flexible framework for optimizing future vaccine-dosing intervals to minimize symptomatic infection risk across populations with diverse immune backgrounds.

## Introduction

Severe acute respiratory syndrome coronavirus 2 (SARS-CoV-2) emerged in 2019 and caused the coronavirus disease 2019 (COVID-19) pandemic worldwide. While the World Health Organization (WHO) announced the end of Public Health Emergency of International Concern (PHEIC), newly infected cases are still emerging as of June 2024, resulting in more than 770 million cases and 7 million cumulative deaths reported worldwide (https://covid19.who.int). Several vaccines have been developed and practically used worldwide to prevent the onset and progression of COVID-19. Among them, COVID-19 mRNA vaccines utilizing the mRNA platform have demonstrated robust protection efficacy by inducing both humoral and cellular immunity. ^1,2^ During the pre-Omicron epidemic period, anti-spike (S) antibody titers against the ancestral strain induced by vaccination were identified as immunological correlates for protection against SARS-CoV-2 infection and severe disease.^3–5^ In contrast, emerging Omicron sublineages repeatedly escaped the immune response elicited by vaccination or prior infection. ^6,7^ While the prevention of severe disease through vaccination was confirmed even during the Omicron BA.1/2 and BA.4/5 epidemic periods, ^8^ a higher anti-S antibody titer against the ancestral strain was required for protection against infection during these periods compared to the pre-Omicron epidemic period. ^9–13^ In addition, humoral immunity induced by vaccination or infection wanes over time, ^14–18^ leading to an increased risk of reinfection. In particular, the residual duration of humoral immunity must be studied to optimize additional vaccination intervals.

Previous studies have reported that individuals who were previously infected, received three vaccination doses, were younger, or were female exhibited higher peak antibody levels than their uninfected, fully vaccinated, older, or male counterparts, respectively. ^14,19^ However, these insights were based on a short-term observation until one-year post infection or vaccination and limited to individuals who received three or less vaccination doses. ^14,19,20^ In Japan, more than 50% of the population received four or more vaccination doses as of April 2023 (Fig. 1a). Additionally, the infection-induced SARS-CoV-2 seroprevalence in Japan was 45.3% for anti-nucleocapsid (N) antibodies during July–August 2023. ^21^ Therefore, models based on these short-term observations and limited vaccination histories are insufficient to predict long-term antibody dynamics in the current, immunologically diverse population. Given the current global and national situations, antibody levels against SARS-CoV-2 are likely diversified due to combinations of multiple vaccine doses, infections with various viral variants, and other background factors. Constructing antibody-waning models considering these diverse factors is crucial for accurately estimating the dynamics of antibody levels in real-world populations. However, this task is challenging due to potential biases in the timing of serum sampling relative to vaccination or infection exposure, often resulting in groups lacking long-term antibody data. For example, by December 2022 in Japan, some individuals who had received their second vaccine dose several hundred days earlier could still be present. Meanwhile, up to five vaccine doses had become available by October 2022 in Japan, with most of the population having received at least three doses, making it difficult to obtain blood samples from individuals vaccinated only twice. Despite these challenges, using a Bayesian hierarchical modelling allows for constructing antibody dynamics models that account for common dynamics across groups and supplement missing antibody levels at specific exposure days for certain subgroups. ^22^ This approach facilitates the estimation of long-term antibody decay dynamics across diverse background populations.

**Fig. 1.**
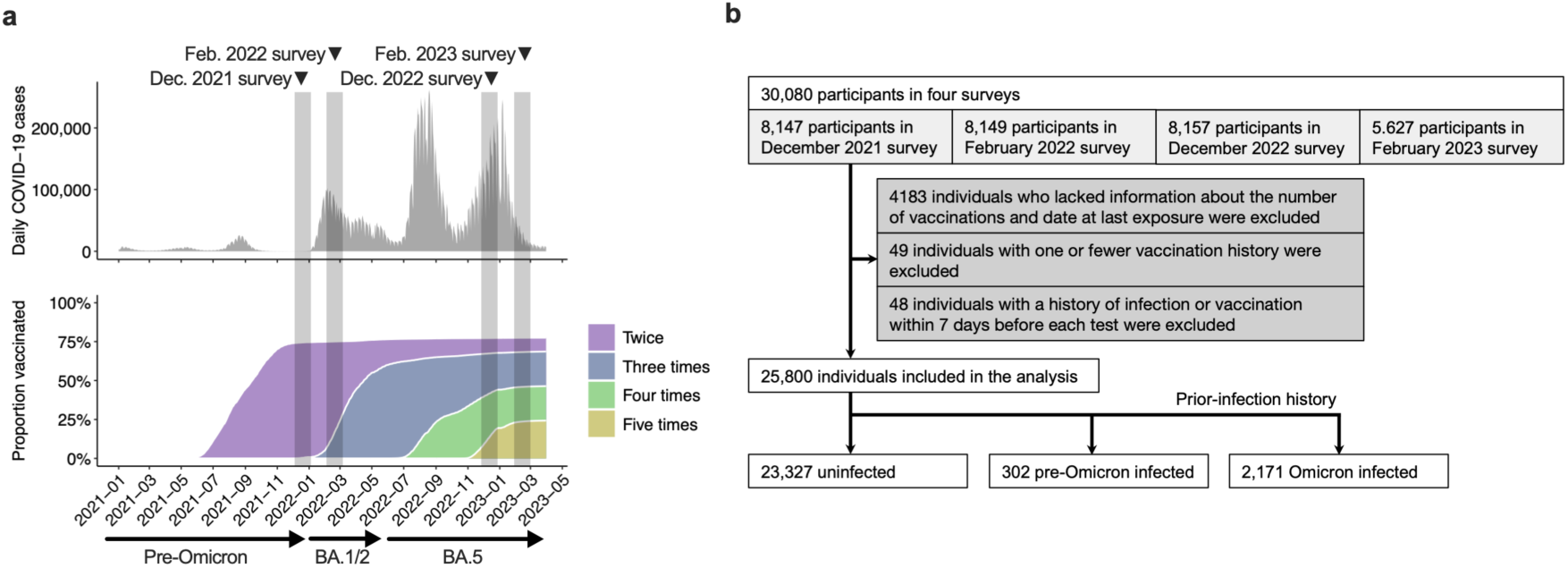
Schematic diagram of this study. (a) Overview of the seroepidemiological survey conducted in Japan for this study. Serum samples and self-administered questionnaires were collected in four rounds conducted in December 2021, February 2022, December 2022, and February 2023. Sampling and data collection were carried out over several days at 15 sites (three municipalities in each of five prefectures). The periods of the surveys correspond to the gray areas in the graph of COVID-19 case counts in Japan (https://www.mhlw.go.jp/stf/covid-19/open-data.html) and the vaccination coverage of the population in Japan (https://www.mhlw.go.jp/stf/seisakunitsuite/bunya/kenkou_iryou/kenkou/kekkaku-kansenshou/yobou-sesshu/syukeihou_00002.html). (b) Flow diagram of the study design and inclusion of participants.

This study aimed to elucidate the long-term serum antibody dynamics across various background populations, such as after multiple SARS-CoV-2 vaccination or previous infection. Using Bayesian hierarchical models, we analyzed anti-S antibody titers and neutralizing antibody measurements from 25,800 serum samples collected from healthy adults during four nationwide seroepidemiological surveys conducted in Japan from December 2021 to March 2023. ^23,24^ We constructed linear and nonlinear models to estimate antibody levels after vaccination or infection over two years to reveal the effects of vaccination history, age, infection history, Omicron variant exposure, and Omicron-adapted bivalent vaccine administration. Furthermore, by integrating these long-term antibody-waning models with established correlates-of-protection models, we aimed to predict the duration of protection against symptomatic infection specific to each immune history. These estimates of long-term protection dynamics, induced by the complex combinations of vaccinations and infections, will provide valuable insights for optimizing future COVID-19 vaccination strategies.

## Results

### Attributes of study participants

We analyzed serum anti-S antibody titers from 25,800 healthy adults in Japan (see Methods, Fig. 1a, b), along with their demographic and clinical characteristics, including age, sex, infection history, vaccination history, and history of Omicron-adapted bivalent vaccination. These data were collected from four nationwide surveys conducted from December 2021 to March 2023 (Table 1). The proportion of individuals with prior infection histories varied across vaccination groups, ranging from 4% to 21%. The proportion of twice-vaccinated participants with prior infection was relatively low, as most were enrolled in 2021 when the seroprevalence of anti-N antibodies was only 2.2% in December 2021, before the onset of the Omicron phase in Japan. ^23^ Furthermore, the seroprevalence of anti-S antibodies in the December 2021 survey was 96.3%, suggesting that almost all participants were fully vaccinated. ^23^ Consequently, we assumed that most participants with a history of prior infection in subsequent surveys represented vaccine breakthrough cases. The interval between last exposure (vaccination or infection) and serum testing among individuals vaccinated twice or three times ranged from 7 to 661 days (Fig. S1).

**Table 1.**
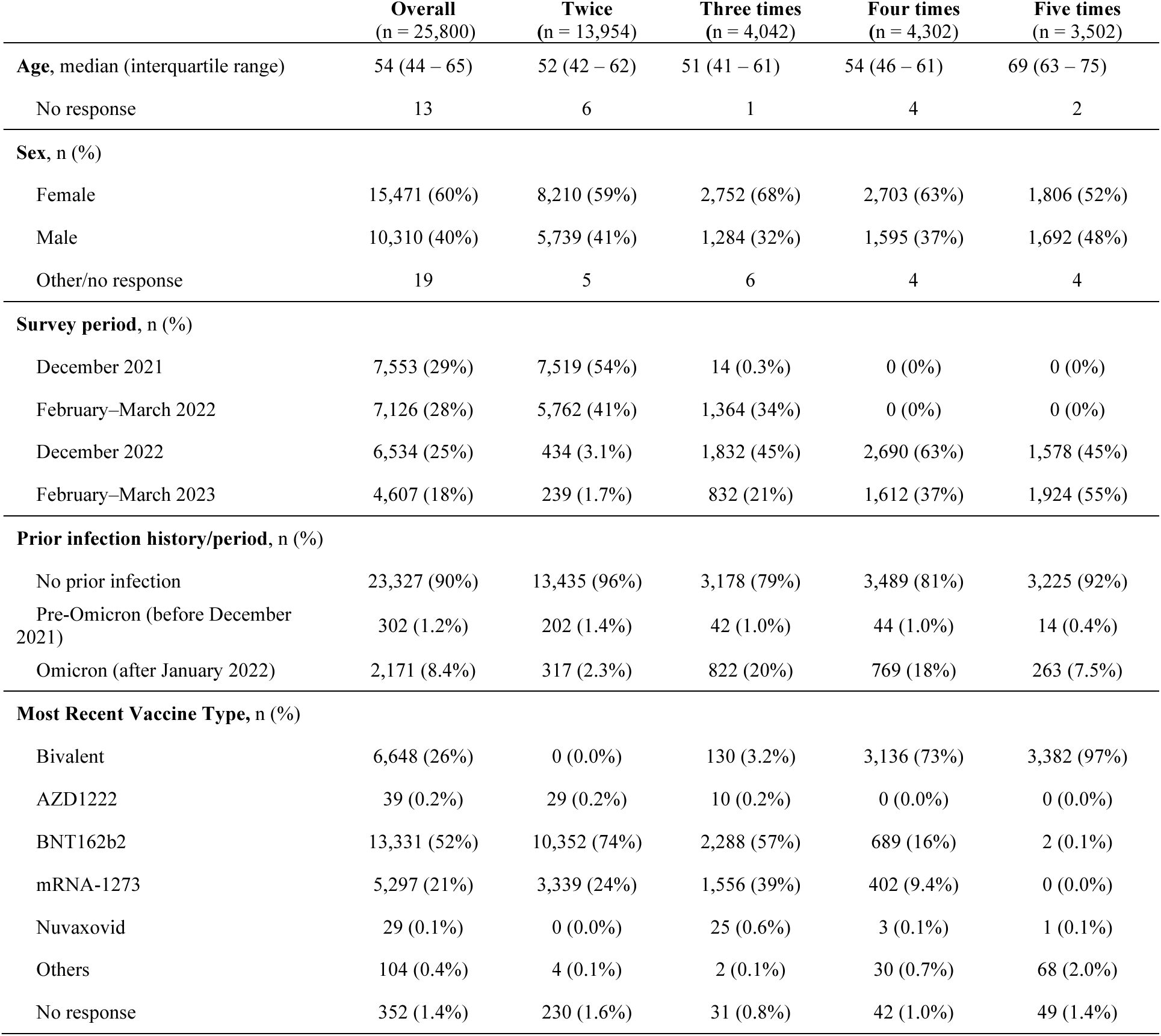
Characteristics of the participants.

Among individuals vaccinated four or five times, 8–18% had documented Omicron infections. Most intervals between the last exposure (vaccination or infection) and serum testing were less than 200 days (Fig. S1), limiting the ability to robustly estimate anti-S antibody kinetics beyond one year using these data alone. Over half of the participants who received four or five vaccine doses had been administered a bivalent vaccine, unlike other subgroups (Table 1). Participants aged 65 years and older were more likely to have received five doses, as the Japanese government prioritized vaccination for this age group (Table 1). For stratified analyses combining vaccination count and prior infection history, the number of samples collected many months after the last exposure was generally smaller within each subgroup. In contrast, sufficient long-term samples were available from individuals vaccinated twice or three times. By constructing hierarchical models using anti-S antibody titers that integrated these subgroups with longer intervals since the last exposure, we were able to estimate antibody titers for subgroups with shorter follow-up periods.

### Estimated kinetics of anti-S antibody titer decay among uninfected individuals with varying numbers of vaccinations

We first evaluated three models to appropriately estimate post-vaccination antibody titer dynamics using serum antibody data from uninfected individuals. Participants were divided into four subgroups based on vaccination count (two, three, four, or five doses) and analyzed using three hierarchical Bayesian regression models. We applied a linear model (M1) and two exponentially decreasing models (M2 and M3) (see Methods) and assessed their goodness of fit. All models demonstrated satisfactory parameter convergence (Gelman–Rubin statistic (Rhat) < 1.01; 50% effective sample size) (Tables S1–S3). Most measured data points fell within the 95% Bayesian prediction intervals of all three models (Figs. 2a–d, S2). However, in twice-vaccinated individuals, the measured values and prediction intervals for M1 and M2 began to diverge beyond 400 days post-vaccination (Fig. S2a, S2f). Bayes factors (BF), calculated using Bayesian free energy via the Bridge sampling method, were used to compare model performance: BF(M2/M1) = 2.5×10^16^, BF(M3/M1) = 1.4×10^79^, and BF(M3/M2) = 5.0×10^61^. These results indicate that the exponentially decreasing model M3, which includes a constant term representing antibody production by long-lived plasma cells, best fit the observed SARS-CoV-2 anti-S antibody titers. Therefore, model M3 was used for subsequent analyses of antibody titer dynamics.

**Fig. 2.**
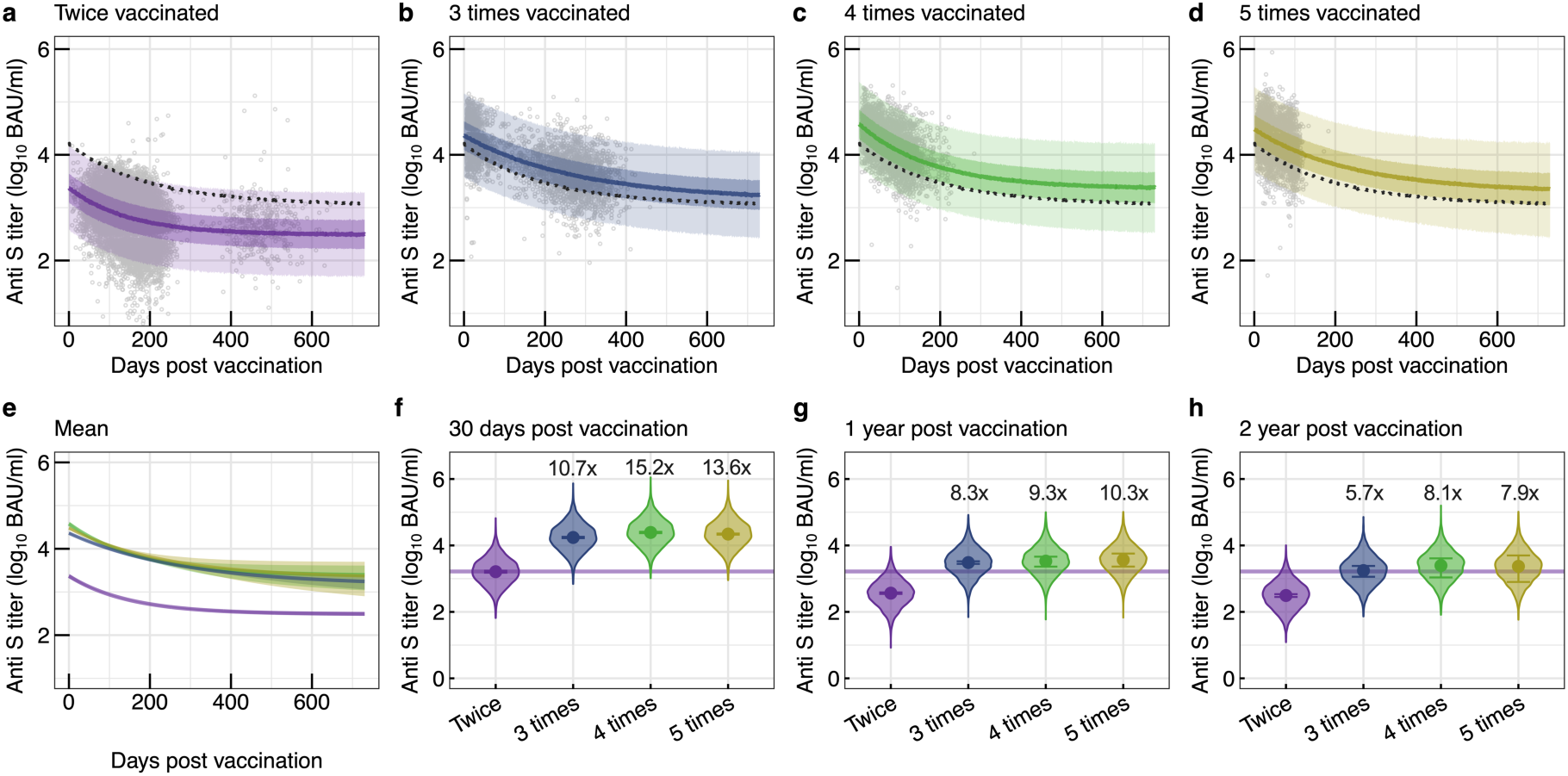
Kinetics of anti-spike (S) antibody titer decay in uninfected individuals estimated with a non-linear (M3) hierarchical Bayesian model. (a–d) Predicted trajectories of anti-S titer decay stratified by the number of vaccine doses, generated with the M3 model. Individual data points are shown as dots. The solid line denotes the median predicted titer, whereas the light and dark-shaded areas indicate the 95 % and 50 % Bayesian prediction intervals, respectively. The dashed line shows the regression curve fitted for all participants. (e) Posterior mean trajectories of anti-S titer decay by vaccine dose. Lines and shaded ribbons represent the posterior median and 95 % credible interval (CredI), respectively. Groups vaccinated twice, three times, four times, and five times are colored purple, blue, green, and yellow, respectively. (f–h) Distribution of anti-S antibody titers by days after vaccination. The 95% predicted distributions of anti-S antibody titers at 30 days, 1 year, and 2 years post-vaccination are shown, along with the posterior distribution median (dots) and the 95 % CredI (error bars). The horizontal purple line indicates the median of the posterior distribution at 30 days after two vaccine doses. Median fold increases against individuals vaccinated twice at the same time point are indicated above the columns (n = 23,327).

Results from the M3 model revealed that anti-S antibody titers over time were higher in individuals who received three or more vaccine doses than in those who received two doses, at 30 days, 1 year, and 2 years post-vaccination (Fig. 2e–h, Table S4). At 2 years post-vaccination, median anti-S antibody titers in the three-or-more-dose group had declined to levels comparable to those observed 30 days after the second dose (Fig. 2h, purple lines). Conversely, the antibody kinetics among individuals vaccinated three or more times were largely overlapping (Fig. 2e–h). These findings suggest that booster vaccinations enhance anti-S antibody kinetics over a two-year period compared with primary vaccination alone but that additional boosters beyond the third dose may not further improve long-term antibody maintenance.

### Age- and sex-related differences in anti-S antibody titer decay

We examined the association between sex and serum antibody titer dynamics (Fig. S3a–c, Table S5). Data from uninfected individuals were grouped by biological sex and further divided according to vaccination count (two, three, or four–five doses), resulting in six subgroups. In this model, both males and females exhibited similar anti-S antibody decay patterns across all vaccination groups (Fig. S3a–c). Furthermore, the estimated antibody titers at 30 days, 1 year, and 2 years post-vaccination differed by less than twofold between males and females (0.8–1.7 fold) (Fig. S3d–f, Table S6). These findings indicate minimal sex-related differences in vaccine-induced antibody kinetics.

We next evaluated the association between age and antibody titer decay (Fig. S3g–i, Table S7). Data from uninfected individuals were stratified by age (<65 or ≥65 years) and further divided by vaccination count, forming six subgroups. Among those vaccinated twice, participants younger than 65 years exhibited 1.7–2.0-fold higher antibody titers at all time points from 0 to 2 years after vaccination (Fig. S3g–i, Table S8). In contrast, smaller age-related differences were observed among the three-dose and four/five-dose groups (Fig. S3j–l, Table S8). These results suggest that while younger individuals initially generate stronger anti-S antibody responses after two doses, booster vaccinations effectively reduce this age-related difference in antibody titers.

### Anti-S antibody titer decay in relation to infection history and Omicron-adapted bivalent vaccination history

We assessed the association between prior SARS-CoV-2 infection history and anti-S antibody titer dynamics (Fig. 3a–c, Table S9). Data were categorized by infection history (uninfected, pre-Omicron, or Omicron infection; see Methods) and further subdivided by vaccination count (two, three, or four–five doses), resulting in nine subgroups. The relationship between days since the last exposure (most recent vaccination or infection) and anti-S antibody kinetics was then estimated.

**Fig. 3.**
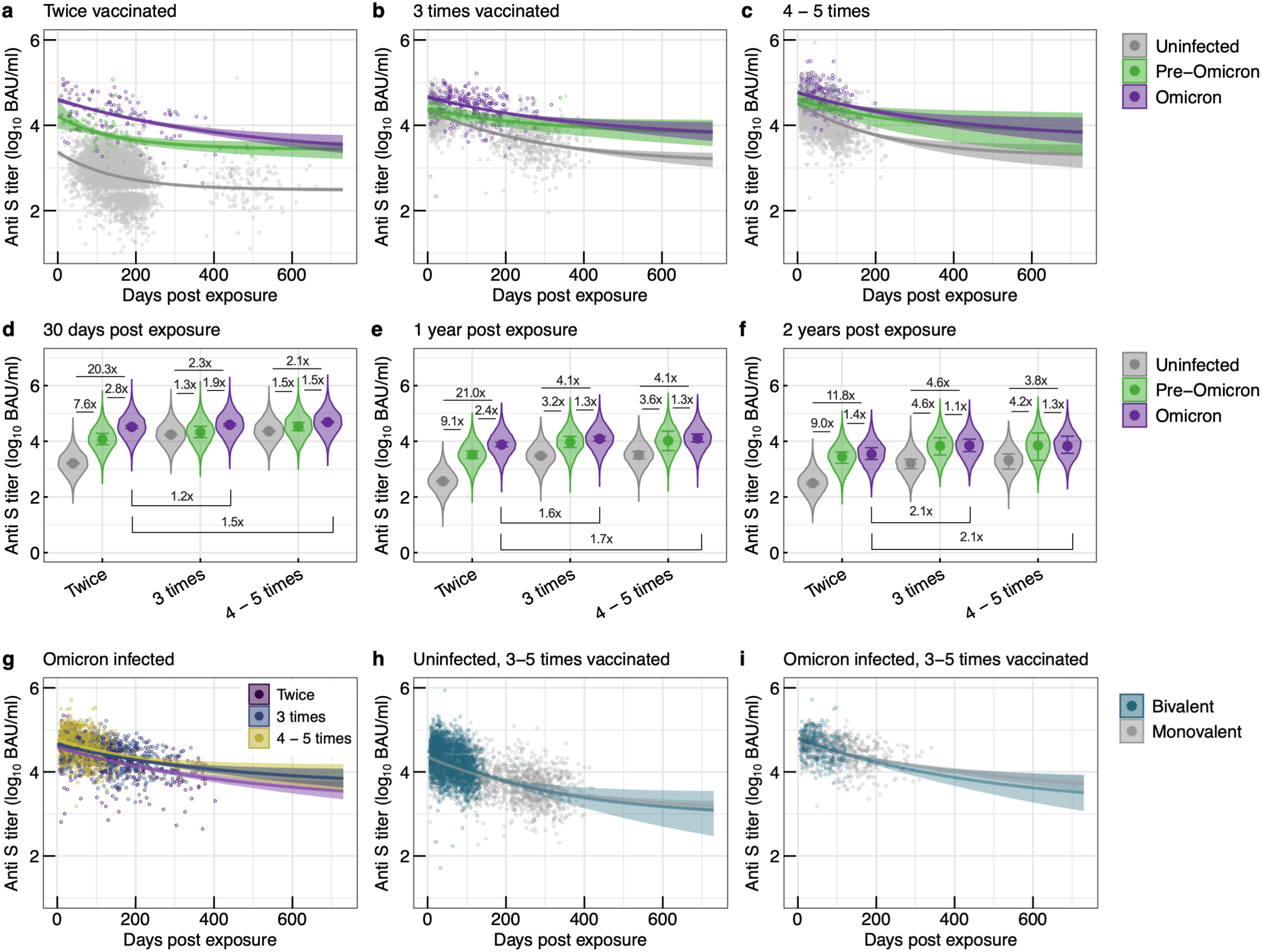
Kinetics of anti-spike (S) antibody titer decay after exposure, stratified by infection history and Omicron-adapted bivalent vaccination history. (a–c) Posterior mean trajectories of anti-S titer decay by combined categories of vaccine-dose number and prior infection status. Solid lines denote posterior medians, and shaded ribbons represent 95 % credible intervals (CredIs). Dots indicate observed titers. (d–f) Posterior predictions of anti-S titers at 30 days, 1 year, and 2 years after vaccination, stratified by infection history. Points show posterior medians; error bars show 95 % CredIs. Median fold-increase values are labelled above (or below) each column. (g) Posterior mean trajectories of anti-S titer decay by vaccine-dose number in individuals with previous Omicron infection (n = 25,800). (h, i) Posterior mean trajectories of anti-S titer decay by Omicron-adapted bivalent vaccination status among uninfected individuals (h) and previously Omicron-infected individuals (i) who had received three to five vaccine doses (n = 11,757).

Among twice-vaccinated individuals, those with prior infection exhibited markedly higher anti-S antibody titers than uninfected individuals (Fig. 3a). Similarly, among participants vaccinated three to five times, prior infection—particularly with the Omicron variant—was associated with higher antibody titers compared with the uninfected group (Fig. 3b–c). In Omicron-infected groups, antibody kinetics overlapped across vaccination counts from two to five doses, suggesting a ceiling effect of breakthrough infection–induced immunity (Fig. 3d–g). Within the twice-vaccinated group, antibody titers up to one year post-exposure followed the order: Omicron-infected > pre-Omicron-infected > uninfected (Fig. 3d–e, Table S10). However, by two years post-exposure, antibody titers among Omicron- and pre-Omicron-infected individuals were comparable, both in the twice-vaccinated group and in those vaccinated three or more times (Fig. 3e–f, Table S10). Moreover, anti-S antibody kinetics after Omicron infection were similar regardless of vaccination count (Fig. 3d–g, Table S10). These findings suggest that SARS-CoV-2 breakthrough infection induces more robust and durable anti-S antibody responses than vaccination alone.

We also examined the effect of Omicron-adapted bivalent vaccination on serum antibody titer dynamics (Fig. 3h–i, Table S11). Data were stratified by infection history (uninfected or Omicron-infected) and further divided by vaccine type (ancestral strain monovalent or Omicron-adapted bivalent), yielding four groups (Fig. 3h–i). The decay patterns of anti-S antibody titers were comparable between bivalent and monovalent vaccine recipients among those vaccinated three to five times (Figs. 3h–i and S4, Table S12).

### Neutralizing antibody decay against Omicron BA.5 in relation to infection history and Omicron-adapted bivalent vaccination

To evaluate the waning of neutralizing antibodies against the Omicron BA.5 lineage—which predominated in Japan during the December 2022 to February 2023 survey period—we performed live-virus neutralization assays in four groups expected to exhibit distinct antibody kinetics: (i) uninfected individuals with two vaccine doses, (ii) uninfected individuals with three to five doses, (iii) Omicron-infected individuals with two doses, and (iv) Omicron-infected individuals with three to five doses. Within each group, serum samples were randomly selected after stratification by the number of days since the most recent exposure (vaccination or infection) in 180-day intervals (see Methods; Fig. S5a). The resulting subset included samples up to approximately 600 days post-exposure in infection-naïve individuals and up to 400 days in Omicron-infected individuals (Fig. S5b). Serum anti-S antibody titers and BA.5 neutralizing antibody titers (BA.5 NT) were strongly correlated (Pearson’s R ≥ 0.80) both within the first year and between one and two years post-exposure (Fig. S6).

Among individuals with two vaccine doses, uninfected participants exhibited almost no detectable BA.5 neutralizing activity throughout the two-year observation period. In contrast, Omicron-infected individuals showed a gradual decline in BA.5 NT, with titers projected to reach approximately 10 NT at around two years post-exposure (Fig. 4a, b). Uninfected individuals with three to five vaccine doses demonstrated mean BA.5 NT values of about 100 immediately after vaccination, which declined below the detection limit within roughly one year. In comparison, individuals with both three to five doses and Omicron infection maintained BA.5 NT values that were 5.3–8.9 times higher than those of uninfected counterparts throughout the two-year period (Fig. 4c–e, Table S13), with mean titers remaining above 10 NT at two years post-exposure.

**Fig. 4.**
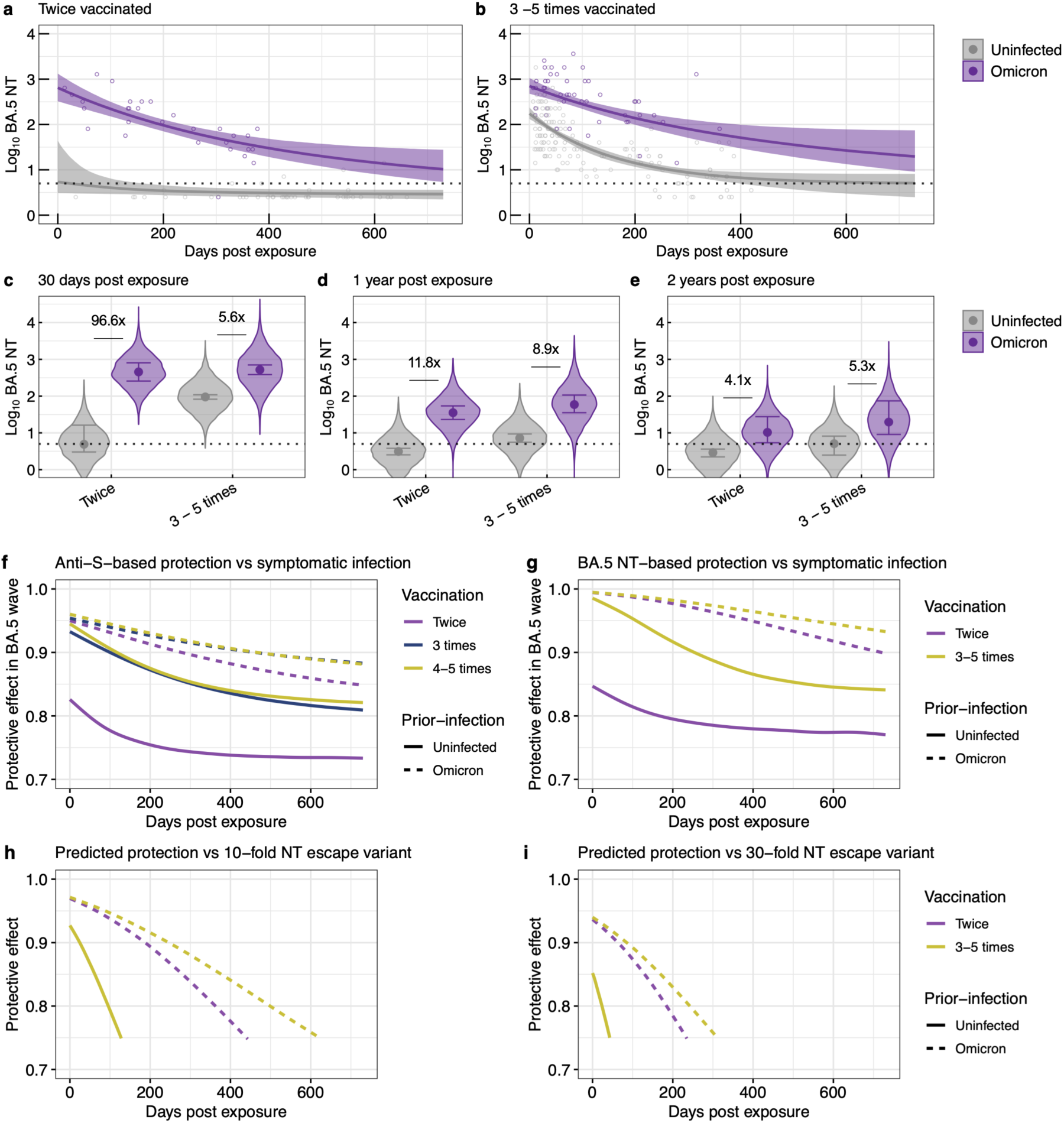
Kinetics of Omicron BA.5 neutralizing antibody titer decay after exposure, stratified by infection history. (a, b) Posterior mean trajectories of Omicron BA.5 neutralizing antibody titer (NT) decay by combined categories of vaccine-dose number and prior infection status. Solid lines denote posterior medians, and shaded ribbons represent 95 % credible intervals (CredIs). Dots indicate observed titers. The black dotted line represents the detection limit (5 NT). The antibody titers below the detection limit were converted to half of the detection limit (2.5 NT). (c–e) Posterior predictions of anti-S titers at 30 days, 1 year, and 2 years after vaccination, stratified by infection history. Points show posterior medians; error bars show 95 % CredIs. Median fold-increase values are labelled above each column (n = 531). (f, g) Estimated protection against symptomatic infection during the Omicron BA.5 epidemic period (December 2022 – February 2023) derived from serum anti-S antibody titers (f) (n = 25,498) and BA.5 NTs (g) (n = 531). (h, i) Predicted protection against symptomatic infection for hypothetical variants that have evaded the BA.5 NTs by 10 times (h) and 30 times (i) based on the serum BA.5 NTs decay model (n = 531). For each group, the posterior mean decay trajectories of BA.5 NTs were converted to protective effect (1 - relative risk), and the resulting median protection over time is plotted.

A parallel analysis of participants who received an Omicron-adapted bivalent booster showed no apparent difference in BA.5 NT decay compared with recipients of the ancestral-strain booster (Fig. S7, Table S14), consistent with findings from short-term (three-month) observational studies. ^25,26^

### Protective period against symptomatic SARS-CoV-2 Omicron infection inferred from serum antibody titers

We previously demonstrated, in a prospective Japanese adult cohort corresponding to the present December 2022 and February 2023 surveys, that serum anti-S titers of 10,209 BAU/mL and Omicron BA.5 NT of 10 confer 90% protection (i.e., 10% relative risk) against symptomatic BA.5 infection. ^24^ Using the antibody-waning models developed in this study, we estimated the decline in protection and the time until titers fell below these thresholds for groups defined by infection and vaccination history (Fig. 4f, g; Table 2).

**Table 2.** Days to reach the antibody-titer threshold for protection against SARS-CoV-2 symptomatic infection during the Omicron BA.5 wave based on serum antibody titer kinetics.

| Infection history | Vaccination history | Days to reach the antibody-titer threshold, Median (95% credible interval) |  |  |  |
| --- | --- | --- | --- | --- | --- |
|  |  | 10,209 Anti-S<br>BAU/mL | 10 BA.5 NT | 10 BA.5 NT<br>(10-fold escape) | 10 BA.5 NT<br>(30-fold escape) |
| Uninfected | Twice | Undetermined | Undetermined | Undetermined | Undetermined |
|  | Three times | 99 (89, 108) |  |  |  |
|  | Four–five times | 116 (111, 121) |  |  |  |
|  | Three–five times |  | 266 (228, 321) | 28 (19, 35) | Undetermined |
| Omicron infected | Twice | 276 (245, 319) | >730 (541, >730) | 195 (154, 238) | 69 (11, 105) |
|  | Three times | 450 (367, >730) |  |  |  |
|  | Four–five times | 450 (343, >730) |  |  |  |
|  | Three–five times |  | >730 (694, >730) | 256 (199, 395) | 92 (63, 123) |

In uninfected individuals, the mean protective duration estimated from the anti-S model was 99 days after three vaccine doses and 116 days after four or five doses—both substantially shorter than the durations observed in individuals with prior Omicron infection (Fig. 4f; Table 2). Among Omicron-infected individuals, even those who had received only two doses were projected to maintain protective levels for 276 days, while those with three or four to five doses were expected to remain protected for approximately 450 days (over one year).

When estimates were based on the BA.5 NT-waning model (Fig. 4a, b), uninfected individuals with three to five doses were expected to retain protection for 266 days (Fig. 4g; Table 2). In contrast, individuals with a history of Omicron infection were predicted to remain protected for more than two years, irrespective of the number of vaccine doses received.

Given that Omicron lineages continue to acquire mutations enhancing humoral immune escape, we further modeled hypothetical variants exhibiting ten-fold and 30-fold reductions in neutralization relative to BA.5 (Fig. 4h, i). Notably, antigenic cartography of plasma collected after breakthrough Omicron infection in individuals primed with an inactivated vaccine revealed that the JN.1 and KP.3 lineages exhibit 15- to 30-fold neutralization escape compared with BA.5. ^27^ For uninfected individuals with three to five doses, the projected protective period against the ten-fold escape variant declined to 28 days, and protection against the 30-fold escape variant was essentially lost immediately after exposure (Table 2).

Even among Omicron-infected groups, the protective duration against the ten-fold and 30-fold escape variants was estimated to be less than one year (Table 2). These findings suggest that if highly immune-evasive variants continue to emerge, booster vaccinations capable of inducing neutralizing activity against circulating strains may be required at least annually.

## Discussion

This study utilized national seroepidemiological survey data from Japanese adults collected between December 2021 and February 2023 to develop a two-year decay model of serum anti-S antibody and Omicron BA.5 neutralizing antibody titers. The model was constructed using hierarchical modeling that incorporated vaccine dose number, infection history, sex, and age groups. Among individuals without prior infection, those who received three or more vaccine doses were estimated to maintain higher anti-S titers over two years compared to those who received only two doses; however, no clear differences in decay dynamics were observed among individuals who received three to five doses. Differences in anti-S antibody decay by sex and age were evident in the two-dose group but were effectively eliminated with three or more vaccine doses. Moreover, a history of Omicron infection contributed to the sustained maintenance of high serum anti-S antibody and Omicron BA.5 neutralizing antibody titers over two years. These decay models, stratified by vaccine and infection history, can flexibly inform optimal vaccine dosing intervals for maintaining low risk of symptomatic infection in populations with diverse immune backgrounds.

Previous studies tracking serum anti-S and neutralizing antibody titers against pre-Omicron strains were generally limited to about one year or included only a few long-term cases. ^20,28^ Our quantitative findings on two-year decay dynamics of serum anti-S and neutralizing antibody titers against Omicron sublineages thus provide important insight for determining the timing of additional vaccine doses. Consistent with prior research, our results indicate that anti-S antibody titers are maintained for approximately one year after two vaccine doses. ^18,20,28^ In this study, long-lived plasma cells contributed to maintaining detectable anti-S titers for two years even among twice-vaccinated individuals. Furthermore, recipients of three or more booster doses exhibited higher anti-S titers than the two-dose group both in the short and long term, maintaining levels for two years equal to or higher than those observed immediately after the second dose.

Studies evaluating the decay dynamics of anti-S titers after four or more doses have so far been limited to approximately six months of follow-up ^20^, leaving real-world decay dynamics unclear. Our findings demonstrate that, among uninfected vaccine recipients, those vaccinated four or five times exhibited anti-S antibody titer increases and decay dynamics equivalent to those of the three-dose group (Fig. 2). This indicates that once robust immune memory is established, additional doses of the same antigen lead to a saturated response. Longitudinal studies have reported an inverse correlation between pre-booster titers and fold increases after the third dose, suggesting that high baseline titers limit the boosting effect of repeated vaccinations with the same antigen. ^29^ This likely reflects antibody feedback, in which memory B cells induced by previous doses are reactivated, reducing the availability of antigen to stimulate new responses. ^29,30^ Nonetheless, our observations suggest that anti-S antibody responses do not diminish over time and that memory B cells responsible for their production remain stably maintained at a saturated level.

Among vaccinated individuals with a history of Omicron infection, both anti-S and Omicron BA.5 neutralizing antibody titers were induced to higher levels and persisted longer than in vaccinated but uninfected individuals. The slower decay observed in Omicron-infected individuals supports the contribution of sustained antibody production by long-lived rather than short-lived plasma cells. Short-term studies have shown robust induction of anti-S and Omicron neutralizing antibody titers in hybrid immunity, particularly after breakthrough infection, ^22,31–33^ and our results quantitatively confirm that these high levels persist for up to two years. While uninfected individuals reached a plateau after the third vaccine dose (Fig. 2), Omicron-infected individuals surpassed this ceiling regardless of vaccine count (Fig. 4). This may reflect distinct routes of immune induction, such as intramuscular vaccination versus mucosal infection, or new antigenic stimulation from the mutated Omicron spike. In Omicron-lineage breakthrough infections, anti-S and neutralizing antibody titers, as well as memory B-cell counts, were markedly elevated. ^32,33^ These memory B cells preferentially recognized epitopes common to both the ancestral and Omicron spikes, suggesting that Omicron infection induces novel antigenic recognition that broadens and strengthens B-cell memory response, sustaining higher antibody titers.

Individuals who received a bivalent vaccine containing the Omicron-lineage spike antigen did not develop anti-S or Omicron-neutralizing antibody titers that were as high or as durable as those observed in Omicron-infected individuals. Short-term studies have reported that the neutralizing and anti-S antibody titers induced by the bivalent vaccine may be comparable to those elicited by the ancestral monovalent vaccine. ^25,26^ This suggests that the ancestral spike antigen component of the bivalent vaccine failed to provide new antigenic stimulation or recognition, thereby limiting the generation of new spike-specific memory B cells. ^34^ Moreover, the viral load at symptom onset in breakthrough infection has been shown to positively correlate with antibody titers during recovery, implying that antigen exposure during infection differs from that during vaccination. ^35^ Collectively, these findings indicate that inclusion of the Omicron antigen in the bivalent vaccine did not enhance the plateaued anti-S antibody dynamics established by prior vaccine doses, likely due to similar administration routes and antigenic properties.

By integrating our antibody-waning models with established correlates-of-protection (90% protection against symptomatic BA.5 infection), we translated long-term antibody kinetics into estimates of protective duration. The projections confirmed the pronounced and durable benefit of hybrid immunity. Based on the BA.5 neutralizing titer model, uninfected individuals with three to five doses were predicted to remain protected for 266 days, whereas those with a history of Omicron infection were expected to maintain protection for over two years, irrespective of vaccine count. This provides a clinical correlate for our earlier observation (Fig. 4): the “saturated” antibody response in uninfected individuals translates to a substantially shorter protection window than the robust, sustained immunity conferred by breakthrough infection. These findings highlight the importance of immune-history-specific vaccination strategies—an uninfected individual with five doses may require a booster much sooner than a two-dose Omicron-infected individual to maintain equivalent protection. However, these projections are based on BA.5. Modeling hypothetical variants with 10- to 30-fold immune escape—similar to emerging lineages such as JN.1 and KP.3—showed that protection declined dramatically. For uninfected individuals, protection against a 30-fold-escape variant was effectively zero. Even among Omicron-infected individuals, the protective period fell to less than one year. These findings indicate that as highly immune-evasive variants continue to emerge, even robust post-infection immunity will wane, underscoring the need for periodic booster vaccinations, likely at least annually, capable of inducing strong neutralizing activity against circulating strains.

This study has some limitations. First, our data were derived from Japanese adults and may not fully generalize to populations with different genetic or healthcare backgrounds. Second, infection history was determined by anti-N antibody status or self-report, which may miss past infections due to waning anti-N responses, leading to possible misclassification. Third, our long-term projections are based on modeling; further longitudinal follow-up is necessary to validate the two-year estimates. Finally, the correlates-of-protection model used was based on data from ancestral or early Omicron variants, and its applicability to emerging variants (e.g., XBB, JN.1) remains uncertain, though relative differences among immune history groups are likely to persist.

In summary, this study demonstrates that additional doses of current vaccines alone do not enhance long-term antibody dynamics against COVID-19, suggesting that the immune response has reached its upper limit. The finding that hybrid immunity from breakthrough infection yields remarkably sustained antibody maintenance suggests qualitative differences in immune responses—such as B-cell durability—not achieved by current boosters. These results support continued development of next-generation vaccines and adjuvants capable of inducing the robust and long-lasting antibody immunity observed after breakthrough infection, thereby informing more effective vaccination strategies.

## Methods

### Survey designs and participants

The dataset used to construct models estimating antibody titer kinetics originated from population-based, serial cross-sectional seroepidemiological surveys conducted in December 2021 (N=8,147), February 2022 (N=8,149), December 2022 (N=8,157), and February 2023 (N=5,627), totaling 30,080 participants. Comprehensive details of these investigations have been previously reported. ^23,24^ Briefly, in each survey, Japanese adults aged 20 years or older from 15 sites (three municipalities within each of five prefectures—Miyagi, Tokyo, Aichi, Osaka, and Fukuoka) were invited to participate. Those who agreed completed a self-administered questionnaire, provided written informed consent, and underwent blood sampling at a designated site. The questionnaire collected demographic data (age, biological sex, occupation, and municipality, among others), comorbidities, COVID-19 vaccination history, and prior COVID-19 diagnosis. Participants in the December 2021, February 2022, and December 2022 surveys were mutually exclusive, while individuals from the February 2023 survey were the same as those enrolled in December 2022.

Participants lacking information on vaccination count or date of last exposure (vaccination or COVID-19 diagnosis) were excluded (4,183 individuals), as were those with one or fewer vaccination events (49 individuals). For antibody decay modeling, 48 individuals tested within seven days of infection or vaccination—during the antibody rise phase—were also excluded. Consequently, 25,800 subjects were included in the analysis (Fig. 1b).

The surveys were conducted as public health investigations under the *Act on the Prevention of Infectious Diseases and Medical Care for Patients with Infectious Diseases* (Infectious Diseases Control Law) and were planned by the Ministry of Health, Labour and Welfare (MHLW) of Japan and the National Institute of Infectious Diseases (NIID). These surveys assessed the prevalence of anti-N and anti-S antibodies, and descriptive results for the full national cohort were published in Japanese on the MHLW/NIID websites. ^36^ The present ad hoc study, conducted as a research activity, evaluated the kinetics of anti-S antibodies using the survey data. Consent for this ad hoc analysis was included in the original survey consent process, which covered agreement for secondary research use of the collected data.

### Ethics

All the samples, protocols, and procedures described herein were approved by the Medical Research Ethics Committee of the NIID and conducted in accordance with the principles of the Declaration of Helsinki (approval numbers 1312, 1457, 1472, and 1730).

### Definition of vaccination and previous infection history

Individuals who reported receiving an Omicron-adapted bivalent vaccine, which includes spike antigens from both the Omicron (BA.1 or BA.5) and ancestral strains, were classified as bivalent vaccinated. Likewise, those who received additional doses of the BNT162b2 or mRNA-1273 vaccines on or after September 21, 2022—the start date of bivalent vaccination in Japan—were also categorized as bivalent vaccinated. Vaccinees who did not meet these criteria were classified as monovalent vaccinated.

Individuals who had been diagnosed with COVID-19 prior to blood sampling were classified as previously infected. To capture unrecognized or asymptomatic infections, participants who tested positive for anti-nucleocapsid (N) antibodies (≥1.0 cutoff index [COI]) were also categorized as previously infected. ^24^ Because the Omicron variant became dominant in Japan after January 2021, participants diagnosed thereafter were designated as Omicron-infected, whereas those diagnosed before this period were classified as pre-Omicron-infected.

### Subsampling for neutralizing antibody titration

To evaluate the decay of NTs against the SARS-CoV-2 Omicron BA.5 variant, we performed stratified random sampling of up to 20 serum samples within each of four post-exposure intervals (7–180, 181–360, 361–540, and 541–720 days) among uninfected and Omicron-infected individuals who had received either two or three to five vaccine doses (Fig. S5). Stored sera from the December 2022 (n = 8,157) and February 2023 (n = 5,627) surveys were screened (total n = 13,784). After excluding 2,628 participants lacking valid information on vaccine dose number or date of last immunological exposure (Fig. 1b), 15 individuals with ≤1 vaccine dose, and none meeting the “<7 days since last exposure” criterion, 11,141 sera remained eligible.

Within each infection group, samples were stratified by vaccine dose number (two vs. three to five doses) and by interval since the most recent exposure (infection or vaccination) into four time windows: 7–180, 181–360, 361–540, and 541–720 days. Up to 20 sera were randomly selected from each exposure-time cell for NT titration, yielding: (1) uninfected, 127 newly selected sera (67 two-dose; 60 three-to-five-dose); and (2) Omicron-infected, 93 newly selected sera (51 two-dose; 42 three-to-five-dose).

The 220 newly selected specimens were combined with the 311 NT results previously reported by Miyamoto et al., ^24^ resulting in a final NT kinetics panel comprising 531 sera. The analytical cohort included 376 uninfected and 155 Omicron-infected samples; pre-Omicron infections were excluded from the decay-model training set due to their limited sample size.

### Electrochemiluminescence immunoassay

Serum samples were heat-inactivated at 56 °C for 30 min before use. Antibody titers for the ancestral spike (S) receptor-binding domain and N were measured using the Elecsys Anti-SARS-CoV-2 S and Elecsys Anti-SARS-CoV-2 kits (Roche, Basel, Switzerland), respectively, according to the manufacturer’s instructions. A COI of 1.0 and a cutoff value of 0.8 BAU/mL were used to determine seropositivity for anti-N and anti-S antibodies, respectively. Because the COVID-19 vaccines approved in Japan are spike-based, anti-N antibodies reflect infection, whereas anti-S antibodies are induced by both infection and vaccination. The numerical results (U/mL) of the Elecsys Anti-SARS-CoV-2 S assay are equivalent to the WHO BAU/mL scale.

### Cells

VeroE6/TMPRSS2 cells (JCRB1819, Japanese Collection of Research Bioresources Cell Bank; Osaka, Japan) were maintained in low-glucose Dulbecco’s modified Eagle’s medium (DMEM; Fujifilm, Osaka, Japan) supplemented with 10% heat-inactivated fetal bovine serum (FBS; (Biowest, Nuaillé, France), 1 mg/mL geneticin (Thermo Fisher Scientific), and 100 U/mL penicillin/streptomycin (Thermo Fisher Scientific) at 37 °C under 5% CO₂.

### Live virus neutralization assay

The SARS-CoV-2 Omicron variant, TY41-702 (lineage BA.5/BE.1, GISAID: EPI_ISL_13241867), was used. Live-virus neutralization assays were performed as described previously. ^37^ Briefly, serum samples were serially diluted two-fold starting from 1:5 in high-glucose DMEM containing 2% FBS and 100 U/mL penicillin/streptomycin and mixed with 100 TCID₅₀ SARS-CoV-2. After 1 h of incubation at 37 °C, the mixtures were added to VeroE6/TMPRSS2 cells seeded in 96-well plates and cultured for 5 days at 37 °C under 5% CO₂. The cells were fixed using 20% formalin (Fujifilm) and stained with crystal violet solution (Sigma-Aldrich, St. Louis, MO, USA). NTs were defined as the geometric mean of the reciprocal of the highest serum dilution that protected ≥50% of cells from cytopathic effect in duplicate wells. Due to limited serum volume, each assay was performed once. All live-virus experiments were conducted in a biosafety level-3 laboratory at the NIID.

### Antibody dynamics model fitting

A Bayesian hierarchical model was employed to estimate the peak and durability of antibody titers. Anti-S binding titers were log₁₀-transformed and treated as the dependent variable (*y*). Participants were categorized into four groups based on the number of vaccinations received (two, three, four, or five doses).

Three models were constructed and validated using antibody titer data from uninfected vaccinees (no history of COVID-19 and anti-N antibody titer <1.0 COI). The first model (M1) was a linear decay model, assuming that log-transformed antibody titers decrease linearly; *y*_[*i*]_ = *a*_[*i*]_ + *b*_[*i*]_x + ε, where *i* denotes the vaccination group, *a* represents the peak titer, *b* the decay rate, *x* the number of days post-vaccination, and *ε* the random error.

The second model (M2) assumed exponential decay: *y*_[*i*]_ = *a*_[*i*]_exp (−*b*_[*i*]_x) + ε, with parameters defined as above. This model reflects the exponential decay of anti-S binding titers.

Since antibody decline after antigen exposure is known to involve both short-lived plasmablasts and long-lived plasma cells, ^38,39^ a third model (M3) was established to reflect this mechanism and is represented by the addition of the term *a*_[*i*]_ exp(−*b*_[*i*]_x), whose value decreases as days post-exposure pass, and the constant term *c*_[*i*]_, which does not change over time; *y*_[*i*]_ = *a*_[*i*]_ exp(−*b*_[*i*]_x) + *c*_[*i*]_ + ε, wherein *a* + *c* represents maximum titers, and *b* represents decay rate of short-lived antibody titers.

In all three models, posterior distributions for each parameter were calculated, with unique values for each vaccination group and common values shared among all groups. These distributions were calculated via Bayesian estimation using the prior distribution of the parameters and the model equation above. Concretely, each prior distribution was set as *a*_[*i*]_∼*N*(µ_*a*_, *σ*_*a*_), *b*_[*i*]_∼*N*(µ_*b*_, *σ*_*b*_), *c*_[*i*]_∼*N*(µ_*c*_, *σ*_*c*_), where *N*(µ, *σ*) denotes a normal distribution with mean μ and standard deviation σ. Additionally, random effect ε adhered to distribution *ε* ∼ *N*(0, *σ*_*y*_); therefore, the titer at time point x is estimable through a normal distribution with mean *y_i_*, and standard deviation *σ_a_*. Weakly informative priors were assigned as normal distributions for *μ_a_*, *μ_b*, *μ_c* and Student’s *t* distributions for the standard deviation parameters (*σ_a_*, *σ_b_*, *σ_c_*, *σ_y_*) (Supplementary Information). Posterior distributions were sampled using Markov Chain Monte Carlo (MCMC) as implemented in *rstan*, with four independent chains of 9,000 iterations (including 1,000 warm-up steps) and thinning every five iterations. Convergence was confirmed for all parameters showed <1.01 Gelman Rubin statistic (R-hat) convergence diagnostic values and effective sample size >1920, i.e., >30%), indicating that the MCMC runs were convergent (Tables S1–3, S5, S7, S9, S11, S15, S16). Model fit was assessed using BF via bridge sampling; a BF(Model₁/Model₂) > 100 indicated strong evidence favoring Model₁.

### Estimation of feature parameters of models

Using the non-linear antibody dynamics Model 3, parameters with distinct values for individual age, sex, infection history, and Omicron-adapted bivalent vaccination history were expressed as *y*_[*i*,*j*]_ = *a*_[*i*,*j*]_ exp(−*b*_[*i*,*j*]_x) + c_[i,j]_ + ε, where *i* denotes the number of vaccination (two, three, or four–five), and *j* represents subgroup characteristics: age (<65 or ≥65 years), sex (female or male), infection history (uninfected, pre-Omicron infection [before December 2021], or Omicron infection [after January 2022]), and Omicron-adapted bivalent vaccination history (bivalent or monovalent vaccination). Posterior distributions for each parameter, with unique values assigned to each group, were estimated using the Bayesian Hierarchal model described above as follows: 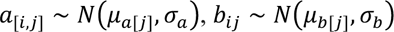, and 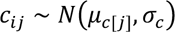. Furthermore, prior distributions for parameters *µ_a_*_[*j*]_, *µ_b_*_[*j*]_, and *µ_c_*_[*j*]_ were also modeled as normal distributions: 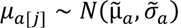, 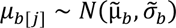, and 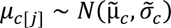. Weakly informed priors were assigned as normal distributions for parameters 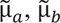, and 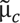 and as Student’s *t* distributions for the scale parameters *σ_a_*, *σ_b_*, *σ_c_*, 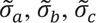, and *σ_y_* (Supplementary Information). Differences between subgroups were evaluated by comparing posterior distributions of model parameters; differences were deemed significant when >97.5% or <2.5% of the posterior mass lay above or below zero. Data lacking sex (n = 18) or age (n = 11) information were excluded from the respective analyses. For analyses involving bivalent vaccination history, data from 13,954 individuals who received only two doses and from 89 pre-Omicron-infected individuals were excluded due to limited representation.

### Infection risk estimation

Risk reduction of SARS-CoV-2 infection during the Omicron BA.5 wave in Japan, based on baseline antibody titers, was derived from our previous longitudinal study using the December 2022 and February 2023 survey cohorts (n = 4,496). ^24^ Briefly, log-transformed antibody titers with cubic spline smoothing were analyzed using Bayesian generalized additive models for newly infected outcomes during the observation period. Models for baseline infection risk were fitted for combinations of anti-N antibody levels with either anti-S titers (Fig. 4f) or BA.5 NT (Fig. 4h–i). Inverse-probability weights for age, sex, survey site, and other demographic factors were applied as previously described. ^24^ The reference risk corresponded to participants with no vaccination and no prior infection (“no exposures”): 16 infections among 87 individuals (absolute risk = 0.184).

For each anti-S titer or BA.5 NT, the conditional absolute infection risk was divided by the reference risk to yield a relative risk (RR), and the protective effect was defined as 1 − RR. Median effect values were plotted against days post-exposure. To model immune-escape scenarios, BA.5 NTs were rescaled by 1/10 (ten-fold escape) or 1/30 (30-fold escape), generating projected protective effect curves for variants with reduced neutralization sensitivity.

## Data Availability

All data produced in the present study are available upon reasonable request to the authors.

## Data Availability

Source data underlying the main figures in this manuscript will be made available prior to publication.

## Code availability

The R codes used for modeling antibody decay will be made available in a Zenodo repository prior to publication.

## Acknowledgments

We thank the Miyagi, Tokyo, Aichi, Osaka, and Fukuoka prefecture governments for their support in implementing the national COVID-19 seroepidemiological survey. We also thank Shoko Sakuraba and Jun Sugihara for their support in implementing the survey from the Ministry of Health, Labour and Welfare, Japan. Additionally, we would like to thank the staff members at the Survey Research Center, Mitsubishi Research Institute, SRL, Inc., and Benefit One Inc. for their administrative and technical assistance with the survey. We thank Chika Furumoto at the Department of Infectious Disease Pathology, National Institute of Infectious Diseases, Japan Institute for Health Security for her technical assistance. The national COVID-19 seroepidemiological survey was funded by the MHLW as a public health investigation. The MHLW funded and was involved in the survey design and selection of the participants for the national COVID-19 seroepidemiological survey to estimate SARS - CoV - 2 seroprevalence in Japan as a public health investigation. The ad hoc study based on the survey data as a research activity was supported in part by a Grant-in-Aid for JSPS Scientific Research (KAKENHI) (23K27422 (to TS) and 23K14534 (to SM)); AMED Research Program on Emerging and Re-emerging Infectious Diseases (JP23fk0108637 (to TS), JP22fk0108509 (to TS), JP 23fk0108684 (to TS), and JP22fk0108568 (to SM)); MHLW Emerging/Reemerging Infectious Diseases and Vaccination Policy Promotion Research Project (24HA2009 (to TS)); Chiba University Futuristic Medical Fund (to SM); Chiba University Synergy Institute for Futuristic Mucosal Vaccine Research and Development (cSIMVa) Vaccine Challenge Grants (to SM).

## Author Contributions

Conceptualization, KN, SM, TS; Methodology, KN, SM; Investigation, KN, SM; Data curation, KN, SM; Formal analysis and Visualization, KN, SM; Funding acquisition, SM, HT, TS; Project administration and supervision, SM, HT, TS; Writing original draft, KN, SM, TS; Writing – review & editing, KN, SM, HT, TS. All authors agreed to submit the manuscript, read and approved the final draft, and take full responsibility for its content including the accuracy of the data and statistical analysis.

