## Supplementary Information for "Predicting long-term waning of anti-spike antibody-dependent protection against COVID-19 across diverse SARS-CoV-2 immune histories"

**Fig. S1–S7 (this file)**

**Table S1–S16 (Excel file)**

**Table S1.** Summary of Model M1 estimates for anti-S antibody titer decay kinetics in uninfected individuals.

**Table S2.** Summary of Model M2 estimates for anti-S antibody titer decay kinetics in uninfected individuals.

**Table S3.** Summary of Model M3 estimates for anti-S antibody titer decay kinetics in uninfected individuals.

**Table S4.** Fold increase in anti-S antibody titers relative to twice-vaccinated individuals among uninfected individuals.

**Table S5.** Summary of model estimates for anti-S antibody titer decay kinetics in uninfected individuals stratified by sex.

**Table S6.** Fold increase in anti-S antibody titers by sex among uninfected vaccine recipients.

**Table S7.** Summary of model estimates for anti-S antibody titer decay kinetics in uninfected individuals stratified by age group.

**Table S8.** Fold increase in anti-S antibody titers by age group among uninfected vaccine recipients.

**Table S9.** Summary of model estimates for anti-S antibody titer decay kinetics stratified by infection history.

**Table S10.** Fold increase in anti-S antibody titers by infection history and vaccination status.

**Table S11.** Summary of model estimates for anti-S antibody titer decay kinetics stratified by Omicron exposure history.

**Table S12.** Fold increase in anti-S antibody titers by Omicron-adapted bivalent vaccination status.

**Table S13.** Fold increase in Omicron BA.5 neutralizing titers by infection history and vaccination status.

**Table S14.** Fold increase in Omicron BA.5 neutralizing titers by Omicron-adapted bivalent vaccination status.

**Table S15.** Summary of model estimates for Omicron BA.5 neutralizing antibody titer decay kinetics stratified by infection history.

**Table S16.** Summary of model estimates for Omicron BA.5 neutralizing antibody titer decay kinetics stratified by Omicron exposure history.

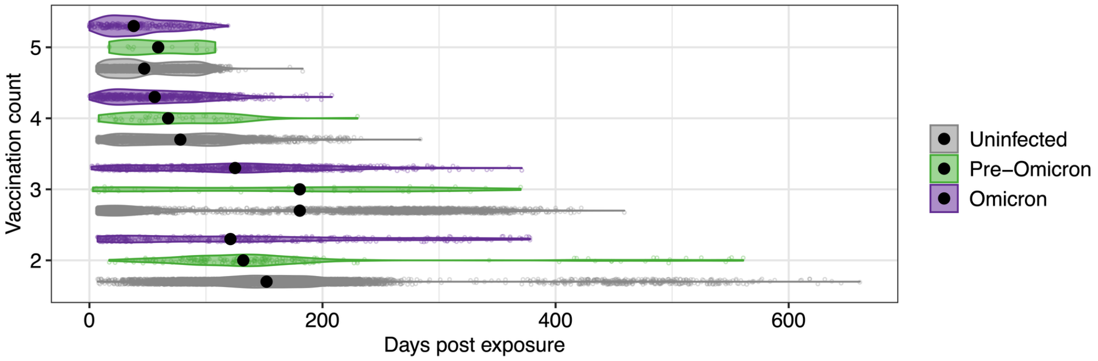


**Fig. S1. Summary of the days from last exposure to serum sampling.**

Days since the most recent exposure, defined as either vaccination or infection, are stratified by the number of vaccine doses and prior infection history, respectively. Each sample appears as a dot, the distributions are displayed as violin plots, and black dots represent the medians (n = 25,800).

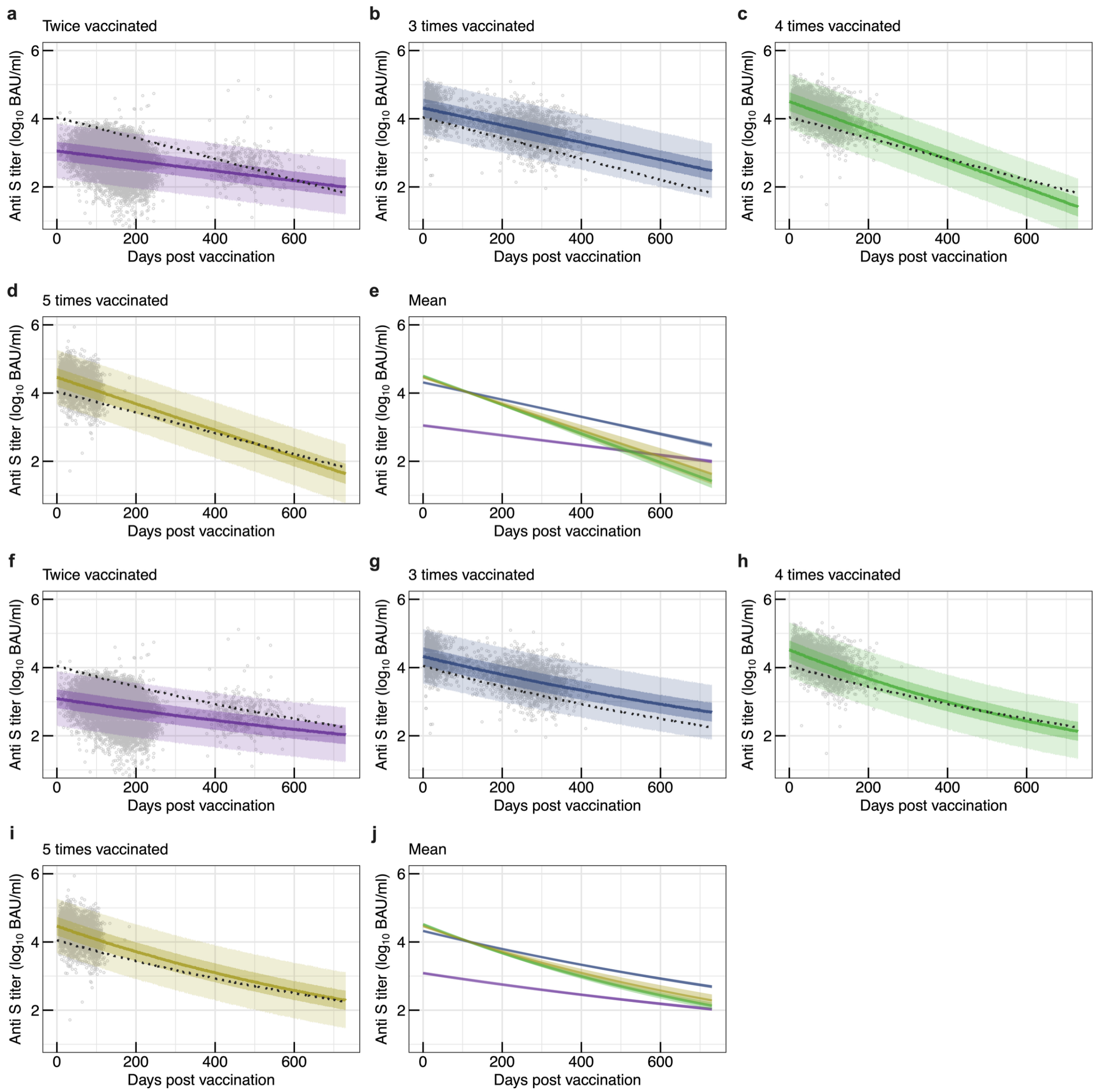


**Fig. S2. Kinetics of anti-spike (S) antibody titer decay in uninfected individuals, estimated using a linear (M1) or nonlinear (M2) hierarchical Bayesian model.**

(a–d, f–i) Predicted trajectories of anti-S titer decay stratified by the number of vaccine doses, generated with the M1 (a–d) or M2 (f–i) model. Individual data points are shown as dots. The solid line denotes the median predicted titer, whereas the light and dark-shaded areas indicate the 95 % and 50 % Bayesian prediction intervals, respectively. The dashed line represents the regression curve fitted to all participants. (e, j) Posterior mean trajectories of anti-S titer decay by vaccine dose, obtained using the M1 (e) or M2 (j) model. Lines and shaded ribbons represent the posterior median and 95 % credible interval (CredI), respectively. Groups of individuals vaccinated twice, three times, four times, and five times are colored purple, blue, green, and yellow, respectively (n = 23,327).

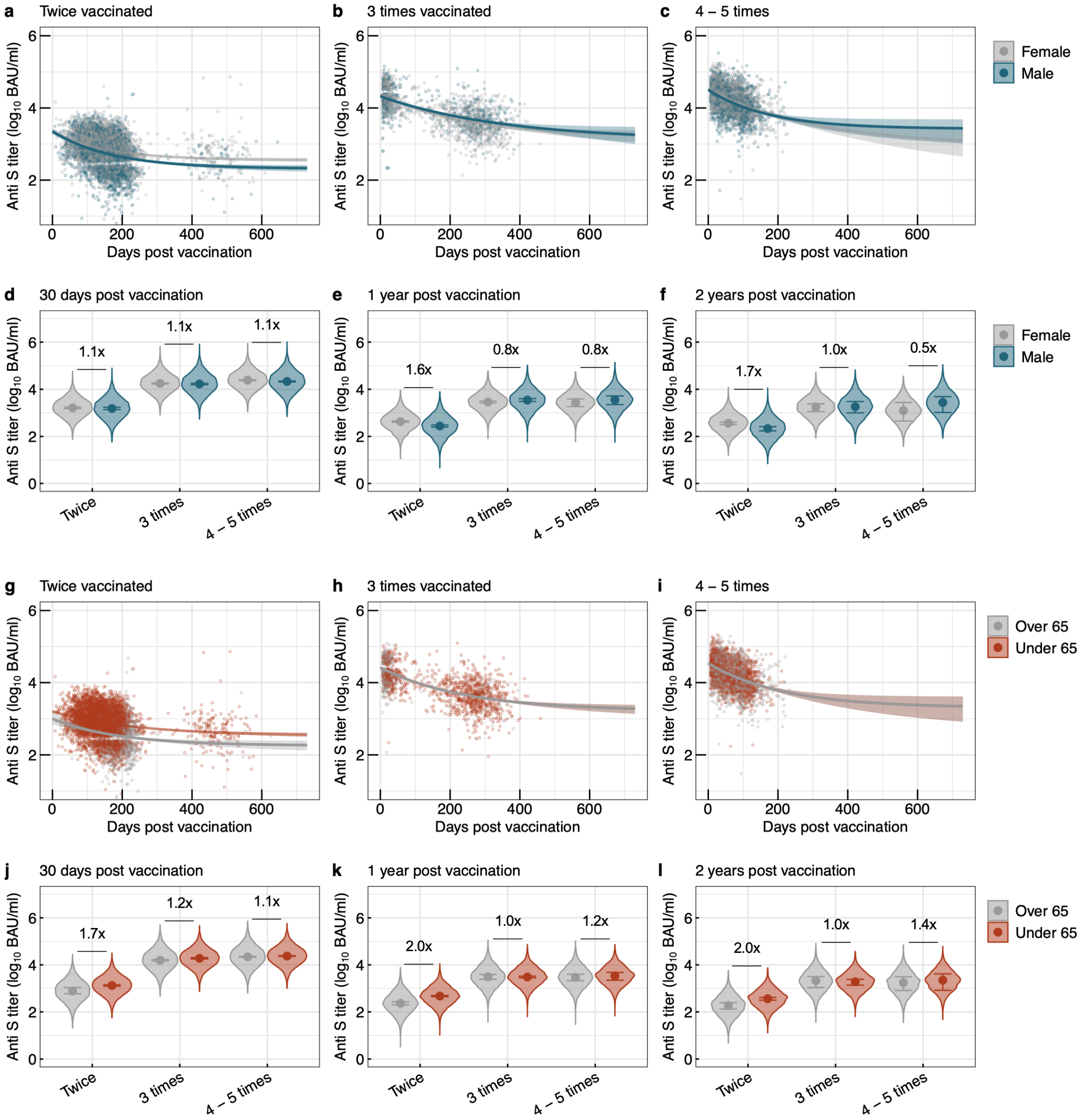


**Fig. S3. Kinetics of anti-spike (S) antibody titer decay after vaccination in uninfected individuals, stratified by sex and age.**

(a–c) Posterior mean trajectories of anti-S antibody decay combined with vaccine dose number and sex group (n = 23,309). Lines and shaded areas represent the median and 95% credible interval (CredI), respectively. Individual data points are shown as dots. (d–f) Sex-specific posterior predictions of anti-S titers at 30 days, 1 year, and 2 years post-vaccination (n = 23,309). Dots indicate posterior medians, and error bars show 95 % CredIs for each prediction. Median fold increases are indicated above the columns. (g–i) Posterior mean trajectories of anti-S antibody decay combined with vaccine dose number and age group (n = 23,316). Lines and shaded areas represent the median and the 95% CredI, respectively. Individual data points are shown as dots. (j–i) Age-specific posterior predictions at the same time points (n = 23,316). Dots indicate posterior medians, and error bars show 95 % CredIs for each prediction. Median fold increases are indicated above the columns.

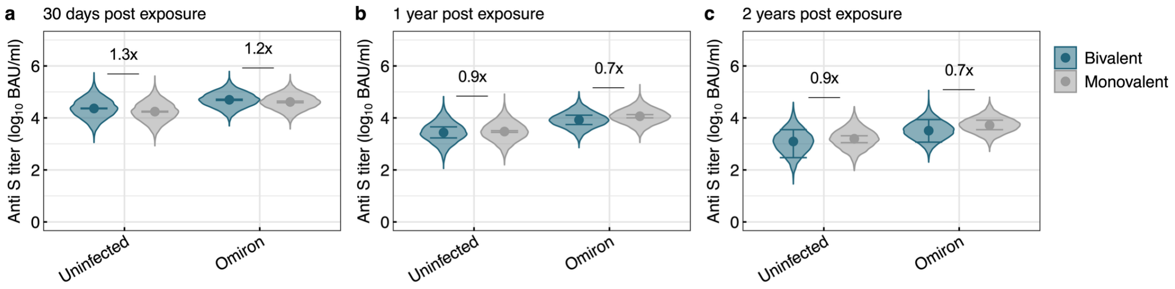


**Fig. S4. Distributions of anti-spike (S) antibody titers at selected time points after vaccination, stratified by infection status and Omicron-adapted bivalent vaccination history.**

(a–c) Posterior predictions of anti-S titers at 30 days, 1 year, and 2 years after vaccination, stratified by infection status and receipt of an Omicron-adapted bivalent vaccine. Dots indicate posterior medians; error bars show 95 % credible intervals (CredIs). Median fold-increase values are labelled above each column (n = 11,757).

a

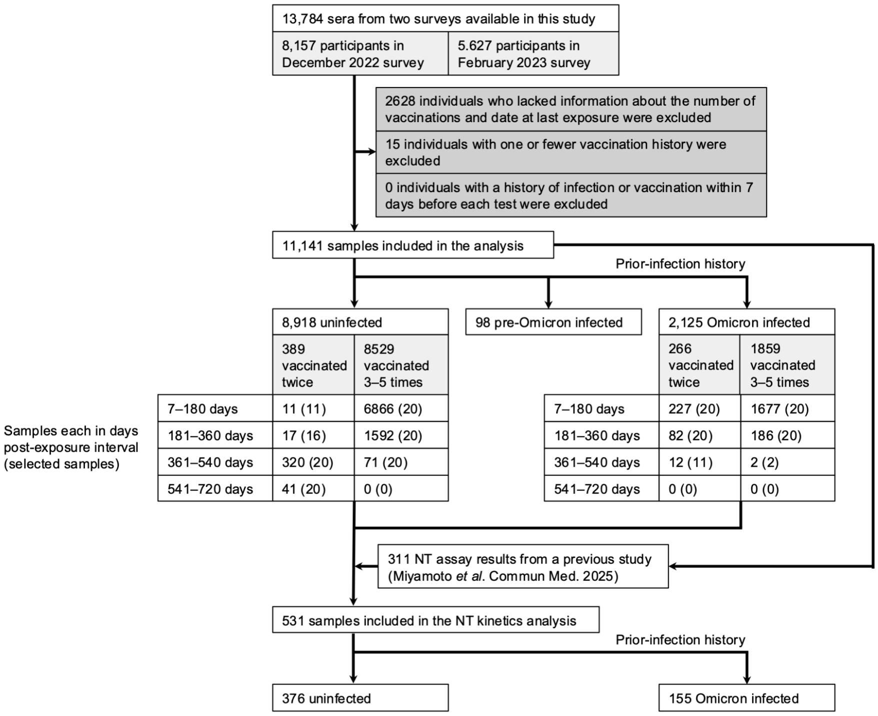


b

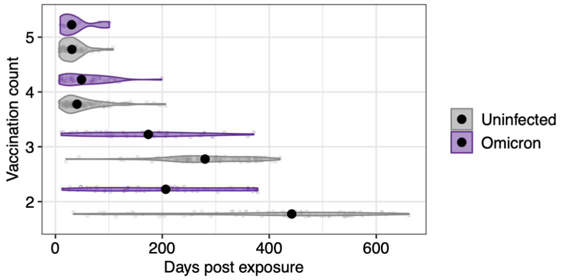


**Fig. S5. Subsampling for neutralizing antibody titer decay analysis.**

(a) Flow diagram of the subsampling. (b) Summary of the days from last exposure to serum sampling. Each sample appears as a dot, the distributions are displayed as violin plots, and black dots represent the medians (n = 531).

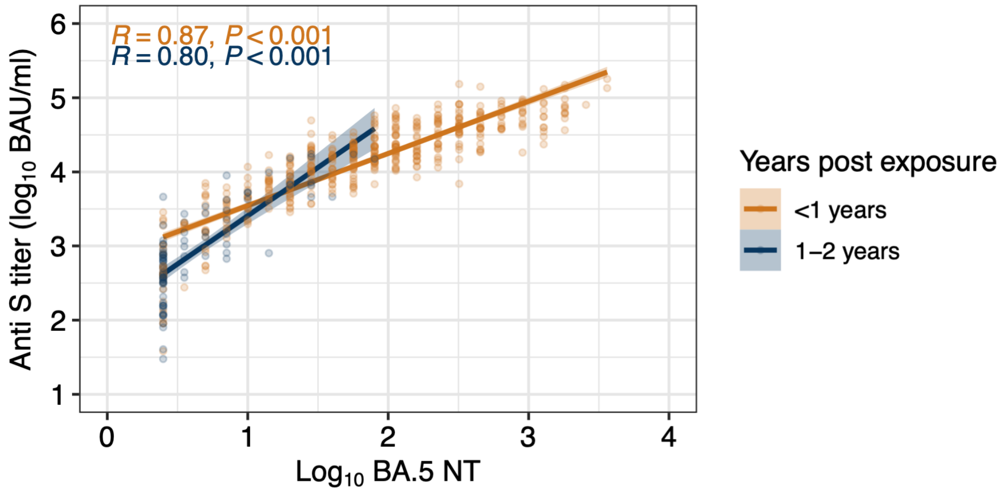


**Fig. S6. Correlation between serum anti-spike (S) titers and BA.5 neutralizing antibody titers (NTs) grouped by years post exposure.**

Data points (blue dots), Pearson correlation coefficient (R), P values, regression line, and 95% confidence interval (ribbon) are shown (n = 531).

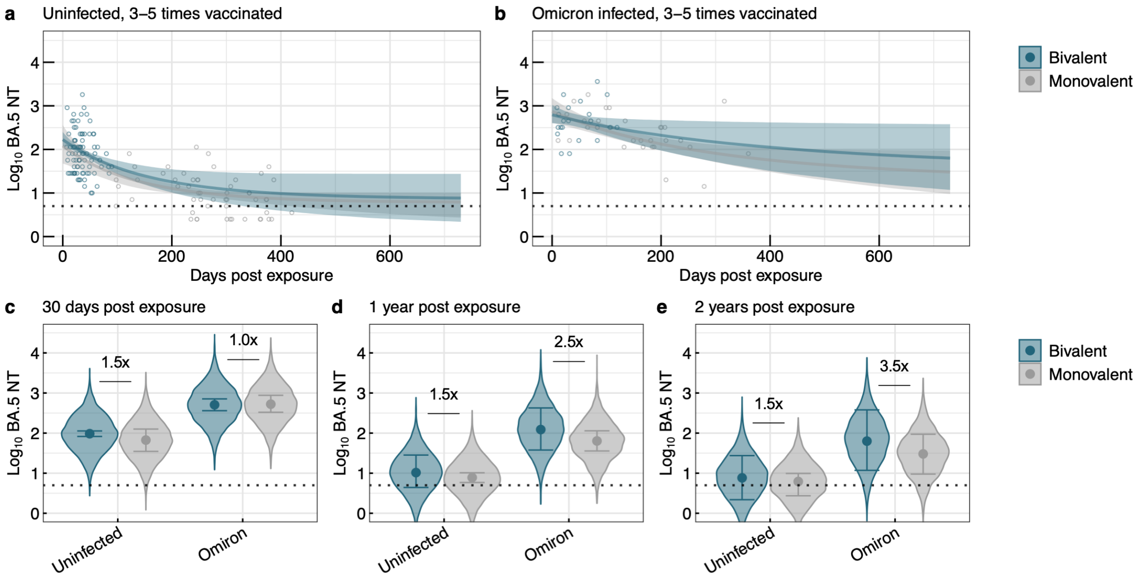


**Fig. S7. Kinetics of Omicron BA.5 neutralizing antibody titer (NT) decay after exposure, stratified by infection and Omicron-adapted bivalent vaccination history.**

(a, b) Posterior mean trajectories of Omicron BA.5 NT decay by Omicron-adapted bivalent vaccination status among uninfected individuals (a) and previously Omicron-infected individuals (b) who had received three to five vaccine doses (n =387). Solid lines denote posterior medians, and shaded ribbons represent 95 % credible intervals (CredIs). Dots indicate observed titers. The black dotted line represents the detection limit (5 NT). The antibody titers below the detection limit were converted to half of the detection limit (2.5 NT). (c–e) Posterior predictions of BA.5 NTs at 30 days, 1 year, and 2 years after exposure. Points represent posterior medians; error bars indicate 95 % CredIs. Median fold-increase values are labelled above each column (n = 387).
